# Breaking Barriers to Universal Health Coverage: Insights from Georgia’s Chronic Disease Medicine Program

**DOI:** 10.1101/2025.02.27.25323038

**Authors:** Alisa Tsuladze, Akaki Zoidze, Nino Kotrikadze, Janina Stauke, George Gotsadze

## Abstract

**Background:** Despite global progress towards Universal Health Coverage (UHC), achieving financial protection remains a challenge, particularly in low- and middle-income countries (LMICs). Out-of-pocket (OOP) payments for pharmaceuticals drive households into poverty and increase inequalities. In Georgia, pharmaceutical costs cause catastrophic health spending, disproportionately affecting the poorest. This study evaluates Georgia’s Chronic Disease Medicine Program (CDMP), explores its evolution and barriers, and proposes strategies to strengthen performance and financial protection.

**Methods:** This exploratory qualitative study combined stakeholder interviews with secondary data analysis. Participants were recruited through purposive and snowball sampling, with semi-structured interviews conducted in person and online. Thematic analysis was conducted to identify key patterns and insights.

**Results:** The CDMP has undergone significant evolution, including expanding benefits and beneficiary groups, adjustments to cost-sharing mechanisms leading to broader population coverage. Recent measures, such as removing limits on reimbursement, have improved participation and service uptake. However, challenges remain: inequitable access, shortcomings in medicine selection, inadequate patient-centered care, limited public awareness, and insufficient involvement of primary healthcare (PHC) providers. While procurement and distribution have improved, capacity constraints and governance issues hinder implementation.

**Conclusions:** The CDMP is a critical step toward achieving UHC objectives and reducing OOP burden imposed by medicine costs. Enhancing the program effectiveness requires prioritizing vulnerable groups, empowering PHC providers, and targeted awareness campaigns. Strengthened governance and increased system capacity are as well critical to overcome remaining barriers and maximize program impact. Findings offer actionable insights for other LMICs seeking to design and implement effective pharmaceutical benefit programs.

## 1. Introduction

Despite global efforts and progress in archiving the Universal health coverage (UHC) agenda to meet sustainable development goals (SDGs), many health systems fail to ensure adequate financial risk protection for their population because of remaining out-of-pocket payments (OOPs) (Bolongaita et al., 2023). Spending on pharmaceuticals and other medical goods is a critical driver of household spending, accounting for 43% of OOPs (global average for 2021) (OECD, 2023). For the poorest consumption quintile, spending on outpatient medicines amounts to 60% of catastrophic health expenditures across 40 countries (Thomson et al., 2023).

The high share of OOP spending on medicines is compounded by epidemiological transition, where non-communicable diseases (NCDs) are becoming the leading causes of death worldwide. In 2016, an estimated 71% of deaths were attributed to NCDs. Approximately 80% of deaths were caused by cancers, cardiovascular diseases, chronic respiratory diseases, and diabetes, with the remaining 20% resulting from other NCDs (NCD Countdown 2030 collaborators, 2018). The importance of NCDs on the global development agenda is well established. The SDG 3.4 aims to reduce premature mortality from NCDs. The SDG Target 3.8 emphasizes the need for achieving the UHC, including access to essential, safe, effective, and affordable medicines and vaccines (Chattu et al., 2023; Wirtz et al., 2017).

Access to medicines (ATM) is important for managing non-communicable conditions as individuals and households affected by NCDs face a higher risk of catastrophic expenditure and impoverishment due to the cost of medications (Kazibwe et al., 2021a). Government-funded benefits play a crucial role in enhancing population health by improving access to essential medicines. It’s important to allocate adequate financing and integrate essential medicines into public sector programs and health insurance schemes (Wirtz et al., 2017) to achieve this. However, improving ATM and ultimately achieving UHC and the SDGs remains a significant challenge for many LMICs (Kazibwe et al., 2021b; Khatib et al., 2016). Therefore, it is necessary to expand the evidence base about LMIC experiences that have attempted to increase access to essential medicines and improve financial protection for the poor while aiming to achieve UHC and SDGs. For this purpose, this paper retrospectively examines Georgia’s experiences since 2017 with the introduction of state-funded drug benefits under the government’s UHC aspirations.

## 2. Background

In Georgia, an upper-middle-income country OOP payments, especially for medicines, have been disproportionately high for decades (The World Bank, 2022). In 2018, medicines accounted for 69% of OOPs, significantly exceeding spending on inpatient (14%) and outpatient care (11%) (Ketevan Goginashvili et al., 2021). According to the latest research in Georgia, the probability of impoverishment increases 43 times for households reporting any expenditure on medicines. These expenses are most problematic for the poorest 20% of the population, where the odds of impoverishment are 45 times higher compared to the wealthiest quintile when all other factors are held constant (Gorgodze et al., 2025).

Chronic illnesses impose a particularly heavy financial burden on Georgian households. NCDs account for 93% of all deaths in Georgia (WHO, 2022). It is expected that the aging population exacerbates this burden further as the share of the population 65 years and older is projected to rise from 16.2% in 2024 to 25% by 2050 (National Statistics Office of Georgia, n.d.; Tbilisi: United Nations Population Fund, n.d.). Long-term treatments for chronic illnesses combined with high costs for medical products are anticipated to significantly increase the financial strain on economically disadvantaged populations. Even small OOP payments for essential medicines can lead to severe economic hardship for this group (Ketevan Goginashvili et al., 2021).

Since 2017, the Georgian government has introduced several pharmaceutical policies to address the financial burden imposed by medicines and improve access to essential pharmaceuticals. Although some programs existed before, such as initiatives targeting oncology medications, diabetes management, and rare diseases, in 2017, the Chronic Disease Medicine Program (CDMP) was introduced to enhance medicine accessibility for patients with chronic conditions. The CDMP provides subsidized access to medications for individuals with chronic conditions and has the highest number of beneficiaries among all state-funded pharmaceutical benefit initiatives. In 2023 the program delivered benefits to 364,545 individuals - surpassing all other programs combined (See Table 1).

**Table 1.** Programs Covering Chronic Diseases.

| State Program | Number of Beneficiaries in 2023 |
| --- | --- |
| Provision of Oncology Medications | 22,476 |
| Palliative Care for Incurable Patients | 3,130 |
| Treatment of Patients with Rare Diseases and Those Requiring Continuous Replacement Therapy | 1,113 |
| Diabetes Management (covers Type 1 diabetes medicines) | 29,669 |
| CDMP (covers 7 conditions: Cardiovascular, Pulmonary, Type 2 diabetes, Thyroid, Parkinson's disease, Epilepsy and Glaucoma) | 364,545 |

All programs listed in Table 1 have specific design, financing and delivery characteristics. Considering that the CDMP has the most extensive coverage and likely the most potential to shoulder drug-related OOPs, we decided to focus our research on this particular program to explore its characteristics, achievements and challenges by answering the following questions: How does the state ensure coverage of medications for the most prevalent health conditions that affect the largest share of the population? What were the program achievements, and what are the remaining challenges? And what measures can help improve the program’s performance?

## 3. Methodology

This study employed an exploratory qualitative research method to understand the design and implementation details of the CDMP and to identify the strengths and weaknesses that could inform potential solutions. For the analysis primary and secondary data sources were used. Primary data were collected through semi-structured interviews, while secondary data were obtained through documentary reviews of legislative documents, administrative data, and relevant published and grey literature.

The research was guided by a thematic analysis organized around the domains of Joose’s 2024 (Joosse et al., 2024) conceptual framework adapted for the study (See Appendix 1). The framework allowed the assessment of core (and relevant) functional areas of the pharmaceutical value chain, which helped describe and analyze the elements of the CDMP design and implementation. The research team developed interview guides with questions focusing on public financing and pricing, selection of medicines included in the benefit package, reimbursement, procurement and supply, healthcare delivery, and dispensing and use. These process elements represented foundational components necessary for understanding the development and implementation of the pharmaceutical benefits program. Together with these process elements, the UHC cube, which helps unpack the benefit package design elements, informs the analysis process (*Principles of Health Benefit Packages*, 2021).

Purposive and snowball sampling were employed to capture diverse expert viewpoints. The initial set of purposefully selected respondents included senior policymakers from the Ministry of Health (MoH) and National Health Agency (NHA, a single national purchaser of health services) and pharmaceutical experts recruited because they were directly involved with CDMP design from the first day. A snowball approach was used by asking initial respondents to suggest the most knowledgeable and informed person with experience and/or a role in the program design, implementation, and/or decision-making.

After securing ethics approval from the Health Research Union’s committee (protocol #2024-01, approved 04/03/2024) and written consent to participate in the research (ensuring confidentiality, voluntary participation, and withdrawal rights), 13 participants were recruited through both means. Interviews were conducted face-to-face meetings at participants’ workplaces and online (via Microsoft Teams or Zoom) in a private setting to maintain confidentiality and minimize distractions. The final sample included current and former MoH senior decision-makers, NHA employees, National Center for Disease Control and Public Health (NCDC) representatives, primary health care (PHC) and pharmaceutical policy experts.

Semi-structured interview guides were developed using the conceptual framework (See Appendix A) and refined based on feedback from the study’s steering committee comprised of health systems, pharmaceutical, and policy experts. These guides were pilot-tested with two participants to confirm the clarity and relevance of asked questions, leading to minor adjustments in question phrasing for improved flow and comprehension. Although no-repeat interviews were conducted, questions were used to clarify emerging themes during the interviews. Interviews were audio- and/or video-recorded and transcribed verbatim. On average, interviews lasted approximately 30-75 minutes. All recordings and transcripts were securely stored on password-protected devices and were accessible only to the research team. During transcription, the respondents’ identifying information was removed to maintain confidentiality. Data saturation was monitored and discussed throughout the data collection process to determine when additional interviews were no longer yielding new information.

Transcripts were analyzed using NVIVO 12™ software. The team employed both deductive and inductive approaches for thematic analysis. Two researchers independently coded the anonymized transcripts to enhance reliability. Discrepancies were resolved through consensus discussions. Regular meetings with the primary investigator ensured consistent feedback and allowed for iterative refinement of emerging themes. The final version of the coding tree is provided in Appendix 2.

## 4. Results

The results section is divided into two main parts. The first one addresses the benefit package design, i.e., the selection process of medicines, beneficiaries covered by the program and medicine cost coverage. This sub-section aligns with the UHC cube, which helps describe the program by providing a structured framework to describe evolutions in coverage dimensions. The Cube’s three axes—services coverage, population coverage, and proportion of costs covered by the program—allow us to visualize the evolution of the benefits package and identify gaps (if any) in access to essential medicines, which are critical for achieving UHC goals (Bigdeli et al., 2015; Roberts et al., 2015). While the UHC Cube provides a valuable framework, it may oversimplify the complexities of reforms when looked at through a health systems lens (Roberts et al., 2015). Therefore, the second part of the results section, as shown on Figure 1, explores the domains influencing the program’s implementation and outputs according to the framework depicted in Appendix 1.

**Figure 1.**
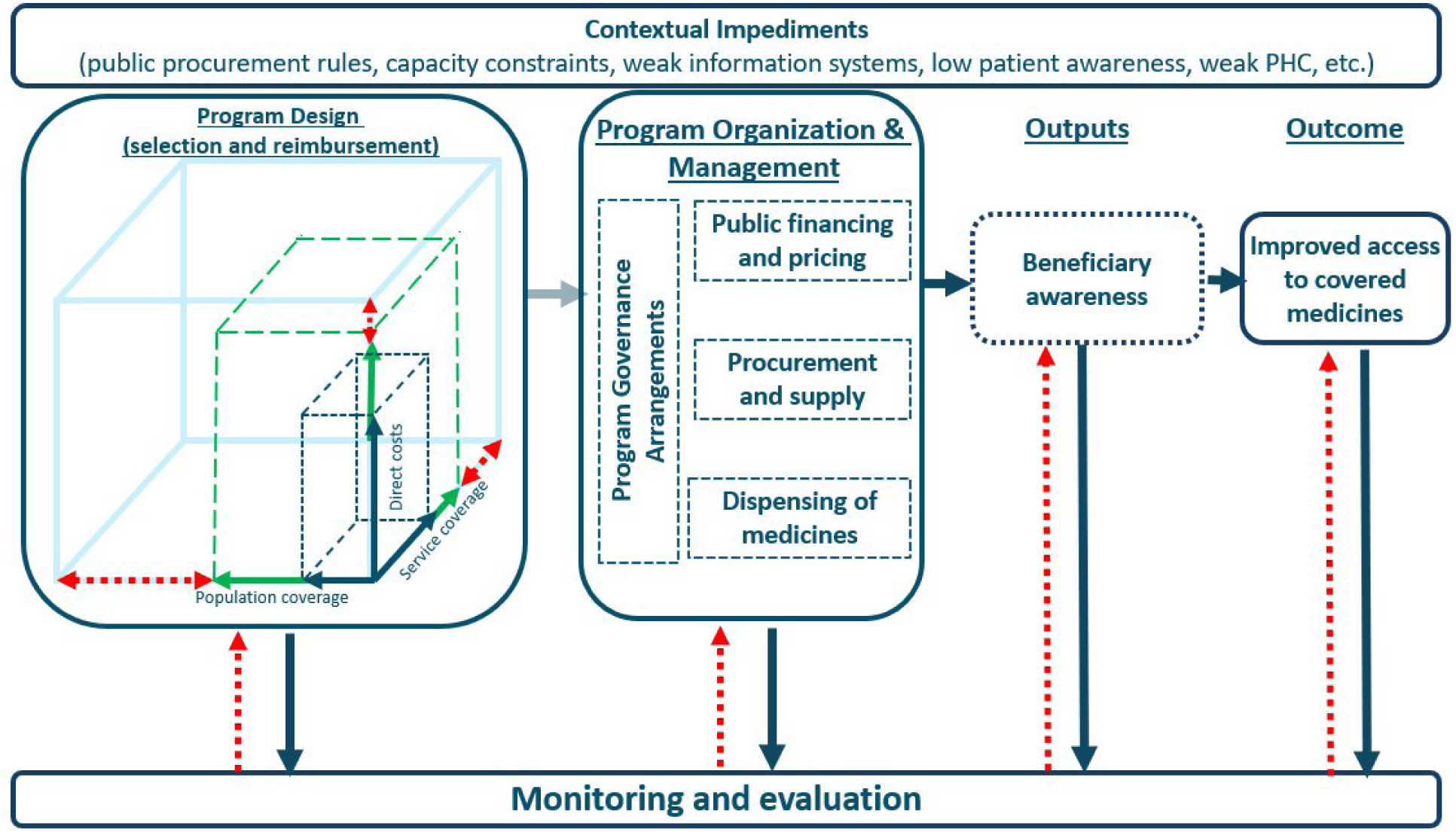
Results Organization Framework.

However, before we delve into the program details, it must be noted that Georgia’s CDMP has been on an evolutionary path since its introduction. Namely, the benefits, beneficiary groups and cost-sharing arrangements have gradually evolved along with program organization, management and implementation arrangements. These developments have positively affected the gradual program expansion and uptake of the benefits. Hence, while describing the study results, we aim to uncover this evolutionary path more granularly. It is essential to understand the reforms over time along with a cross-sectional analysis to reveal factors determining success or remaining challenges.

### 4.1 Program design

#### 4.1.1 Service coverage

At the program’s inception, key stakeholders—including the MoH, the NCDC, USAID’s Health Improvement Project experts, and other specialists—used data from the STEPS survey (a Step-Wise approach to NCD Surveillance) (NCDC, 2010) to identify chronic diseases with high prevalence contributing to high mortality rates for inclusion in the program.

> “The projections were made based on the most prevalent chronic diseases. All specialty experts were involved, and we agreed that we needed to fund diseases with the highest burden and mortality rates” (respondent 7).

In 2017, the program began by selecting health conditions - cardiovascular diseases, type 2 diabetes, chronic obstructive pulmonary disease, and chronic thyroid disease. Later Parkinson’s, epilepsy (2019) and glaucoma (2024) were added. This approach aimed to ensure that medicine coverage was provided for the most prevalent and, thus, priority health conditions.

In 2017, a critical step in the program’s design was selecting medicines. Key stakeholders, including the MoH, NCDC, and USAID experts, collaborated to determine the required medicines and quantities needed to treat the selected conditions. Demand calculations targeted 60% of eligible beneficiaries, accounting for anticipated program uptake, with quantities based on annual dosage requirements.

Significant challenges emerged in aligning the medicine list with national clinical guidelines, which often recommended combination therapies. Instead, the program prioritized single-component drugs for their lower price, driven by a cost-minimization strategy mandated by public procurement rules. This approach prevented the inclusion of combination medicines in the program and likely contributed to its low uptake (State Audit Office of Georgia, 2020).

Several respondents highlighted patient dissatisfaction with the program’s treatment options, noting that the limited coverage failed to meet their clinical needs or expectations. This disconnect between patient preferences and the program’s offerings emerged as a key barrier to its success during the initial years:

> “When adding medicines, the Ministry employs a cost-minimization approach. For instance, when introducing a new molecule, it is assumed that different forms of the medication are bioequivalent. Consequently, the Ministry selects the three least expensive forms available on the market.” (Respondent 13)

> “The main reason patients dislike this program is because of the types of medicines it includes. The medications aren’t effective [in patients’ opinion], which is why patients drop out of the program.” (Respondent 3b)

Thereafter, the modifications were introduced to the medicine list requested by patients or different associations. These requests were managed by the NHA Committee^1^. The amendment process included consultations with professional associations, pharmaceutical experts, and primary healthcare doctors to improve the diversity of recommended medicines. Although the stakeholders actively participated in this process, the MoH retained the final decision-making authority, limiting the committee’s autonomy and initiative. Consequently, this structure may have led to a bureaucratic process in which political priorities overshadowed technical considerations/rationale:

> “Perhaps the functioning of the committee is more formal than professional, depending on what the political [not technical] priority is at the moment.” (Respondent 12)

Incorporation of patient-centeredness in medicine selection proved to be limited, with financial considerations frequently taking precedence over clinical needs/preferences. Despite the involvement of experts, there was scant evidence that patient or doctor preferences were duly considered, raising concerns about the program’s capacity to address public health needs (demands) effectively.

#### 4.1.2 Population and cost coverage – reimbursement

Initially, the program focused on socially vulnerable groups in 2017, later expanding to include pensioners, children with disabilities, and elderly individuals with significant disabilities in 2018.

> “Initially, the program was intended solely for the socially vulnerable. However, since we aimed to cover chronic conditions, we also included pensioners [because of the high prevalence of selected NCD among people above 65 years]. The target number of beneficiaries was set at 300,000 people, encompassing both the socially vulnerable and pensioners.” (respondent 11)

The program’s initial design until 2024 included annual reimbursement limits per patient and condition determined by subject matter experts and professional associations. The amounts for limits were derived from rough projections of annual patient consumption of medications and the lowest prices available on the market. However, the evidence and process through which the final limits were determined were often criticized as unclear decision-making, as they consequently proved to be grossly inadequate, frequently leading to mid-year or even earlier exhaustion of allocated amounts. In many cases, these limits failed to meet the real needs of patients with chronic illnesses, such as diabetes and cardiovascular conditions.

> “Somewhere in the middle of the year, the limit for cardiovascular conditions, for example, was exhausted, and one after another, letters came from the patients. One [letter] said: ‘let’s sleep in the summer and wake up in the winter’ [when the limits are renewed at the beginning of the calendar year].” (Respondent 3a)

Over time, as it became evident that the existing limits were not adequately addressing patients’ needs, policymakers were prompted to reassess the situation. After years of debate and negotiation, particularly with the Ministry of Finance (MoF), policymakers implemented the reform in 2024, culminating in abolishing the monetary annual limits for medications.

> “We worked for several years to remove this limit on medicines, we faced a lot of opposition from the Ministry of Finance, but finally we reached an agreement and the limits were removed.” (Respondent 3a)

This shift represented a significant change in the program, driven by the recognition that the management of chronic diseases cannot be effectively achieved with restrictive blanket caps on reimbursed amounts. Removing the limits allows patients to access the medications they need throughout the year without constraints.

### 4.2 Program Organization and Management

#### 4.2.1 Program Governance Arrangements

As mentioned in the previous section, the committee established to make relevant decisions and changes in the program design functioned largely as a formality, often regarded as a mere tick mark rather than an active governing body. The committee failed to provide strategic direction and ensure that the program’s design evolved based on evidence and needs. Without an effectively functioning committee, decision-making remained superficial, limiting the program’s ability to timely adapt and improve.

The State Audit (State Audit Office of Georgia, 2020) revealed significant gaps in oversight, particularly in the monitoring of storage conditions and inventory management (in the early years of the program). While the agency conducted warehouse inspections and performed inventory checks, responsibilities were poorly divided. The same individuals were tasked with both receiving medications and verifying their presence, creating a conflict of interest that undermined accountability.

A fundamental principle of governance is transparency (World Health Organization, 2007), yet several critical aspects of the program lack it. The budgeting process and the selection of medicines at the program’s inception were not conducted with sufficient openness, limiting stakeholder confidence in these decisions. Additionally, while the NHA publishes reports on program utilization, these reports are neither comprehensive nor easily interpretable by the general public.

The accountability mechanisms within the program are significantly weakened by the absence of an effective Monitoring and Evaluation (M&E) system. The State Audit Report highlighted the lack of comprehensive performance indicators, making it difficult for the MoH to assess progress and enforce accountability among departments and individuals involved. Without clear benchmarks and evaluation metrics, as well as clearly defined responsibilities, there is no structured way to ensure program goals are being met.

#### 4.2.2 Public Financing and Pricing

Public financing of medicines has evolved throughout the years, particularly in budget determination and allocation. Since 2017, the CDMP has operated as a separate budget sub-program under the MoH, managing the procurement and distribution of medicines. The budget was primarily based on selected health conditions, the list of selected medicines, their estimated quantities and their respective market prices.

> “The NCDC provided projected beneficiary numbers, while the estimated costs for chosen medicines came from the National Health Agency. The expected budget requirement was calculated by multiplying these two values” (Respondent 13).

An analysis of NHA statistics from 2017 to 2020 revealed a low budget uptake for the program during this period), a finding further corroborated by the state audit report (State Audit Office of Georgia, 2020) and later confirmed by respondents, who noted that the program had limited reach.

> “Over the years, there has been low consumption [of medicines].” (Respondent 3a)

> “Ministry bought half [medicines quantities] of what experts predicted, and despite this, because of low patient referrals, drug spending grew very slowly.” (Respondent 11)

Starting in 2020, the CDMP was integrated into the Universal Health Coverage Program (UHCP). The CDMP sub-program budget was eliminated, and its financial needs were embedded within the larger UHCP budget allocation. This shift enabled flexible resource allocation and promoted data-driven decision-making and interdepartmental collaboration. It moved beyond sole reliance on the MoH’s financial department, which had limited capacity for planning and managing sub-program finances.

> “There were times when only the accountants did the work [the budget estimation],” while currently, “different departments are actively involved [in the planning-budgeting process]” (Respondent 11).

This evolution in participation has been instrumental in gathering critical data on costs, monitoring medicine uptake and budget spending patterns, and enabling the removal of program limits in 2024 after reviewing prior years’ medicine consumption trends. These changes were reinforced by introducing an external reference pricing (ERP) policy in 2023 and enhancements in the information system, which allowed the MoH to oversee imported quantities of medicines, distribution, consumption, national stock levels and monitoring prices across the supply chain. These improvements have strengthened logistics oversight, increased pricing transparency, and optimized stock management.

> “When we started reference prices as a control mechanism, we developed a digital stock management system that manages the stock and monitors the import of medicine into the country, where [geographically] it is sold, and at what price. It’s the whole cycle [that is monitored] until it [medicine] reaches the patient” (respondent 2).

Despite observed progress, challenges persist in budget estimation. There is a gap in the technical expertise required for budget preparation, notably lacking the health economics expertise:

> “How can a public finance accountant prepare a health budget without knowing epidemiology and health economics?” (Respondent 12).

> “In the area of health economics, the ministry has limited potential and therefore relies on external experts and their support” (Respondent 11),

Such expertise was only made available earlier in the process (around 2017) through US-funded technical assistance, but since MoH was not able to tap into an expert pool. Integrating the chronic medicine program into UHCP reduced the need for the MoH to produce precise budget estimates and defend it before the MoF, addressing capacity limitations evident in earlier program design.

Furthermore, the budget estimation process lacks established methodological rigor and predominantly relies on budget expenditure tracking, again highlighting institutional capacity weaknesses. This reliance on historical data rather than systematic analysis and forecasting is evident, as one respondent states:

> “Our budget mainly relies on the previous year’s expenditure” (Respondent 11).

Respondents also pointed out that much of the budgeting process remains non-transparent because the internal calculations that account for unit prices and required quantities (based on expert opinions or budget expenditure data) are not standardized into formal methodological guidance. The absence of official guidance fosters performance inconsistencies on an individual level:

> “… internal documents never became a formal technical document, which could methodologically guide consistent conduct of employees involved in the process [of budget preparation]” (Respondent 11).

Therefore, there is certain distrust in the process and its outcomes, and the decisions appear to be ad hoc:

> “The process of defining the budget was not entirely clear … eventually, how it was finally decided and what was [the budget] allocated is still unclear to me “(Respondent 7).

In conclusion, key informants point out that broader fiscal policies adopted for UHCP primarily shape budget determination and allocation in healthcare, including for drugs.

#### 4.2.3 Procurement and supply

Initially, during 2017-2019, the national procurement arrangements for the CDMP were managed by the MoH, which oversaw both the procurement and distribution/logistics through contracted entities. However, the program faced significant challenges in quantifying the required medicines, timely conducting public procurement tenders, and distributing drugs from the central store to selected/contracted regional pharmacies, leading to frequent shortages that impeded access. As one respondent noted:

> “The ministry purchased these medicines; however, some expired, some we could not procure, and others we had in stock but were limited in quantities. When a patient came to the pharmacy with a prescription, they might receive one of the prescribed medicines, but for the others, the pharmacy would say they were unavailable, and the patient would be asked to return later. This was a significant problem for this program” (Respondent 3).

> “My patient visits the pharmacy, and then I receive a call from them that the pharmacy is suggesting an alternative medication to the one I prescribed. When our doctors visit the pharmacy to check, they find that the commercial brand is available, but the state-funded medication is out of stock.” (Respondent 7)

The centralized procurement system managed by the MoH struggled to align supply with actual demand, leading to surpluses in some geographies and shortages in others, exacerbated by variable regional demand. Additionally, the tendering process faced challenges, such as a lack of suppliers in 2018. In 2019, consolidated tenders were introduced to streamline procurement, but they caused further complications, with many medicines remaining unavailable. As respondent N4 explained,

> “Procurement started with consolidated tenders in 2019, and due to the transition to this model, they had difficulty making purchases. Many medicines could not be bought, leading to shortages.”

At first, only five pharmacies in Tbilisi and one in each municipality^2^ participated in the program, limiting geographical access, especially for rural residents. This also highlighted the need for a more extensive pharmacy network.

Recognizing the described challenges, the MoH implemented new arrangements in 2020, exploiting the specificity of the pharmaceutical market structure of Georgia, which has a well-established retail pharmacy footprint throughout the country owned and operated by the private sector. Furthermore, several vertically integrated pharmacy chains manage pharmaceutical import, wholesale and retail sales in the country. This allowed MoH to contract these private chains for procurement, storage, and distribution, ensuring drug availability nationwide. As a result, starting in 2020, patients could access medicines at any pharmacy of their choice, and pharmacies could dispense drugs using an e-prescription platform and be reimbursed by the NHA monthly.

> “The methodology changed the [program] administration itself. Until 2020, the agency conducted procurement and carried out distribution in pharmacies, and patients went there and received it [medicine] from there. The agency was responsible for logistics. Since 2020, the model has changed, and the patient went to the pharmacy chains and received these specific medicines there” (respondent 4).

#### 4.2.4 Dispensing of medicines

While engagement of the private pharmaceutical sector in CDMP helped resolve many procurement, logistical and patient access problems observed during the early years, these benefits came with some risks. In Georgia, recent studies have revealed collusion between providers and marketing practices used by pharmaceutical companies when, because of financial benefits, treating doctors often steer patients towards higher-cost medications (Tvaliashvili et al., 2024), even in cases when affordable state-funded alternatives are available. Therefore, providers and/or pharmacies sometimes “fail” to inform patients of cheaper government-funded alternatives. Or pharmacists could mislead patients, claiming that these medications are not currently unavailable and promoting more expensive options for which patient has to pay out-of-pocket.

> “Patients were informed by the pharmacy that the state-funded medication was unavailable, but a commercial alternative was available for purchase.” (Respondent 6)

In 2022, the MoH issued a regulation mandating doctors to prescribe medicines with generic names via e-prescriptions^3^ and pharmacists to offer patients up to three alternatives of the prescribed medicine at the lowest price available in stock as a remedial action for described pharmacy malpractice. Although the recent State Audit Report states that 67% of the medicines is dispensed without e-prescription (State Audit Office of Georgia, 2024).

During the program’s early phases, patients faced significant administrative hurdles that hindered access to state-funded medicines. To qualify for benefits, individuals had to visit a territorial unit of the NHA and complete a registration process. This process required presenting an identity card and submitting an official patient summary (Form N100) provided by their treating doctor. The form confirmed the diagnosis of a chronic disease (using ICD-10 codes) and included details of the prescribed medication and daily dosage. Any changes in the prescription mandated the re-submission of an updated Form N100 to renew registration. Only after completing these steps could beneficiaries access medications at designated pharmacies. This cumbersome, multi-step administrative procedure—requiring visits to multiple locations with corresponding travel costs—often led to significant delays, posing a substantial administrative barrier to timely treatment.

> “Administration was challenging. Patients needed to obtain a prescription, visit the NHA, wait for approval, and then go to the pharmacy for their medication. For something [benefit] that cost only 1 to 3 GEL, patients had to go through such a lengthy process and spend so much on travel that the medicine’s value was almost irrelevant. This complexity has kept uptake low over the years.” (Respondent 3)

In response to these challenges, administrative and bureaucratic barriers were significantly reduced after the program was integrated into the UHCP in 2020, and accessibility was enhanced.

### 4.3 Monitoring and evaluation

Since its early days, the CDMP’s M&E has significantly evolved. In 2020, the State Audit Office of Georgia identified critical areas for improvement in M&E. The report stated, *” M&E indicators are inadequate to ensure the program is effective and attaining its goals. Beneficiaries are not uptaking procured medicines, which means the program can’t achieve accessibility objectives.”* Additionally, the report revealed that internal control mechanisms were inadequate for managing medicine shelf life and preventing waste (State Audit Office of Georgia, 2020).

In response, the MoH introduced a more robust pharmaceutical management information system, as described earlier. This system enables the MoH to monitor the entire supply chain of medicines, from importation to patient dispensing. Private pharmaceutical companies manage inventory in accordance with Good Distribution Practice standards, while the MoH’s system tracks geographic distribution, pricing, and availability across the supply chain.

Using this information system, MoH’s Policy and Strategic Development and Analytics Departments collaboratively conduct routine data analysis. When issues emerge, the NHA is alerted to address the problem promptly. Thus, routine monitoring of medicine consumption, program utilization, and patient turnover ensures the smoother program implementation:

> “Drug consumption, utilization, and patient visits are tracked—not just beneficiary registration. We also monitor whether the process is continuous, checking how many times they [patients] took the medicine, whether it was just once or ongoing” (respondent 3).

Despite these improvements, challenges persist, primarily stemming from the information system’s inadequate design. The system fails to produce standardized analytical dashboards and necessitates considerable time and effort for routine analysis, hindering thorough monitoring and evaluation efforts within an environment characterized by limited human resource capacity at the ministry or its subordinated units.

Removing the limits on pharmaceutical benefits signified a positive development, but it also introduced new challenges, particularly in prescription monitoring and budget controls. Eliminating these limits has sparked concerns regarding the potential for physicians’ overprescription, which could strain the state’s healthcare budget in 2024. According to respondents, the risks resulting from limit removal are managed with the help of an enhanced monitoring system, which should help respond to unexpected changes in utilization. The Strategic Development and Analytical Department of MoH plays a key role in this effort, closely tracking and analyzing significant changes in prescription patterns to identify and address potential over-prescription, thereby helping safeguard the budget. However, managing this risk depends more on the individual’s capacity within the department and less on systemic solutions (e.g., dashboards routinely generated from management information systems, etc.). On the contrary, the data generation to be used in the analysis remains time-consuming and requires significant effort from NHA and MoH staff.

### 4.4 Beneficiary Awareness

Interviews also uncovered low user awareness about state-funded benefits among the public as a significant contributing factor to the low program uptake, especially observed during 2017-2019 (State Audit Office of Georgia, 2020). While the government made strides in raising public awareness through television ads, posters, and social media campaigns, these initiatives took place during the program’s first year. Some respondents noted that most of the population still lacked knowledge of the program and its benefits.

> “Lack of information about the benefits” has contributed to the program’s low uptake (Respondent 1).

> “The communication has been less frequent this year [2024] than in the previous period… Unfortunately, chronic conditions seem less publicized than other medicines-related policy changes, including reference-pricing and oncology medicines program” (Respondent 2).

Periodic television briefings and press conferences, intended as the primary sources of information about CDMP benefits, were insufficient to achieve lasting public awareness. A 2019 Ministry-commissioned survey confirmed low levels of program awareness among the population, with only 13% of respondents reporting familiarity with program details (State Audit Office of Georgia, 2020). The Ministry’s efforts in 2017–2018 to stimulate demand for the program appeared inadequate, as notable referral growth was only observed after the active campaign launched in late 2019, which reached 40% of potential beneficiaries.

Finally, primary care providers, who should lead in informing patients about the program benefits and prescribing medicines, were not actively involved, partly due to limited knowledge or adverse motivations (described earlier). Public skepticism toward government programs further contributed to patient reluctance. Additionally, some patients perceived government-supplied medicines as inferior due to their lower price, associating higher-cost medicines with better quality.

> “It is noteworthy that the population does not believe in the government promise until the doctor explains it and prescribes it to the population [the medicine covered under the program].” (Respondent 13)

### 4.5 Program Outcomes

As noted earlier, the CDMP had many design and implementation challenges initially, leading to low uptake and program benefits. Gradually, the numerous adjustments made to program design, revised implementation arrangements for logistics and distribution, and revisions in medicine selection and co-payment policies helped increase the program outcomes. Consequently, the number of beneficiaries increased to 390,555 by the end of 2024 (or 14% of the adult population), and the program expenditure grew from 0.5 in 2017 to 58 million GEL in 2024 (or 116 times).

## 5. Discussion and Recommendations

This study highlights the significant progress and persistent challenges of Georgia’s CDMP in advancing UHC to reduce the burdening costs of medicines the population pays on an OOP basis. While notable improvements have been achieved in population, service, and cost coverage, the program’s implementation has been constrained by design flaws, governance limitations, and capacity shortfalls. These findings provide critical insights for improving the CDMP and offer valuable lessons for other low- and middle-income countries (LMICs) seeking to enhance access to essential medicines under the UHC agenda. Before delving into the details, one critical insight emerges from this work. While political will and commitment to providing NCD medicines under a UHC scheme are essential, they are far from sufficient. Without a well-functioning and resilient health system—where all its components are effectively aligned and adequately resourced—the scheme will struggle to deliver its intended benefits. Therefore, timely ensuring seamless procurement, adequate supply chain management tailored to a country context, adequate financing, workforce capacity, and service delivery is crucial for translating policy ambitions into meaningful health outcomes (Healthy Systems for Universal Health Coverage, 2017).

### Key Achievements

Since its inception, the CDMP has evolved in scope, gradually expanding beneficiary groups and service coverage, which took almost eight years. Removing reimbursement caps in 2024 marked a pivotal improvement, facilitating easy financial access to medicines and addressing one of the program’s key design weaknesses – insufficient coverage of medicines due to inadequately established reimbursement limits based on expert opinions and not on actual consumption data. Next, leveraging Georgia’s extensive private pharmacy network instead of centrally planned procurement and distribution has simplified program logistics and enhanced geographic accessibility, significantly mitigating early challenges in medicine distribution and availability. Georgia’s dense private pharmacy network—145 pharmacies per 100,000 population, compared to 28 in OECD countries (*Final Report on the Monitoring of the Pharmaceutical Market, Obtained Results, and Recommendations Issued by the Agency*, 2022; Kane, 2023)—provided a conducive context for such decisions and allowed the MoH to delegate procurement and distribution responsibilities to the private sector, which, over the decades, developed strong capacity in handling logistics for pharmaceutical distribution. With the support of the European Union, Georgia has legislated *Good Distribution Practice* (GDP) standards for pharmaceuticals (Tabatadze et al., 2024) and many companies have already met the requirements by obtaining certification from external certifying bodies. Consequently, effective public-private collaboration improved service delivery, demonstrating how such partnerships can address logistical challenges faced by MoH through effective collaboration and mutual accountability, as shown also elsewhere (Abdul et al., 2024).

Achievements in information system development need to be noted. Initially, when the CDMP was conceptualized, necessary data for evidence-based decision-making was completely lacking. The data about medicine prices was not readily available, decision-makers had no idea about regional consumption patterns, and stock management was challenged. All of this initially hampered the program design and implementation. Nonetheless, reforms, such as the introduction of an ERP policy in 2023, which also triggered enhancements of the Health Information System (HIS), have improved pricing availability and transparency, effective stock management, and monitoring of medicine consumption patterns on a patient level. Many of these developments were also achieved through public-private collaboration and data exchange between private and public players involved in the program implementation, obviously supported by necessary regulations and governance arrangements.

Administrative barriers constraining patient access to benefits during the early days of CDMP were eventually reduced/removed. Complex registration and benefit renewal processes imposed significant challenges, and beneficiaries questioned the value of pursuing limited benefits versus significant time and transportation cost investments. This has fostered skepticism among the population about the program’s ability to improve access to medicines and their health outcomes. However, the eventual removal/reduction of these barriers most likely contributed to growth in benefit uptake, as seen in Figure 2. These achievements underscore the program’s potential to reduce OOP expenditures further and improve financial protection for covered populations (especially poor and vulnerable) if and when benefits are further expanded and the depth of coverage is increased.

**Figure 2.**
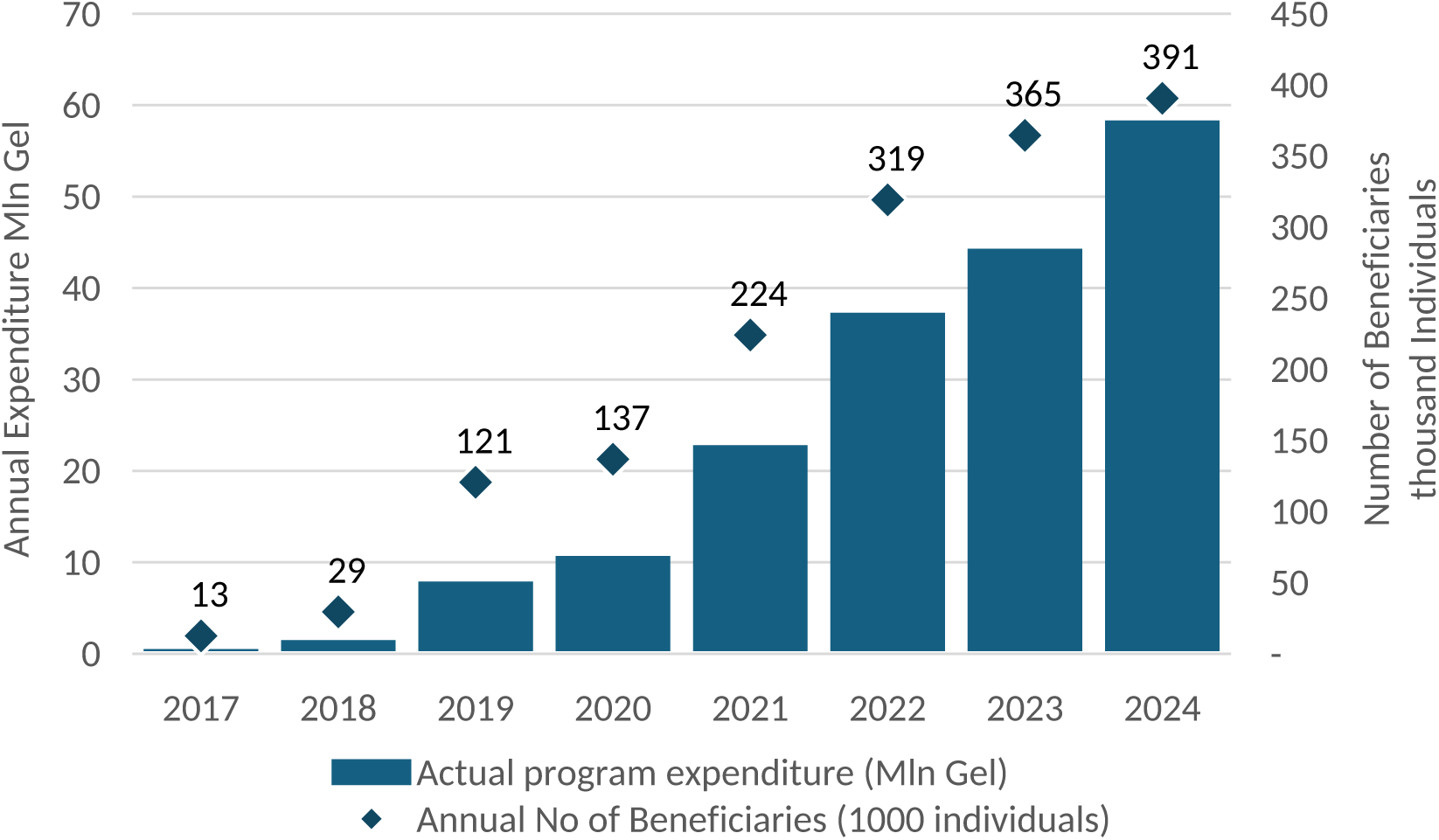
Annual Chronic Disease Medicine Program Expenditure and number of beneficiaries 2017-2024. Source: https://nha.moh.gov.ge/ge/c/statics

**Figure 3.**
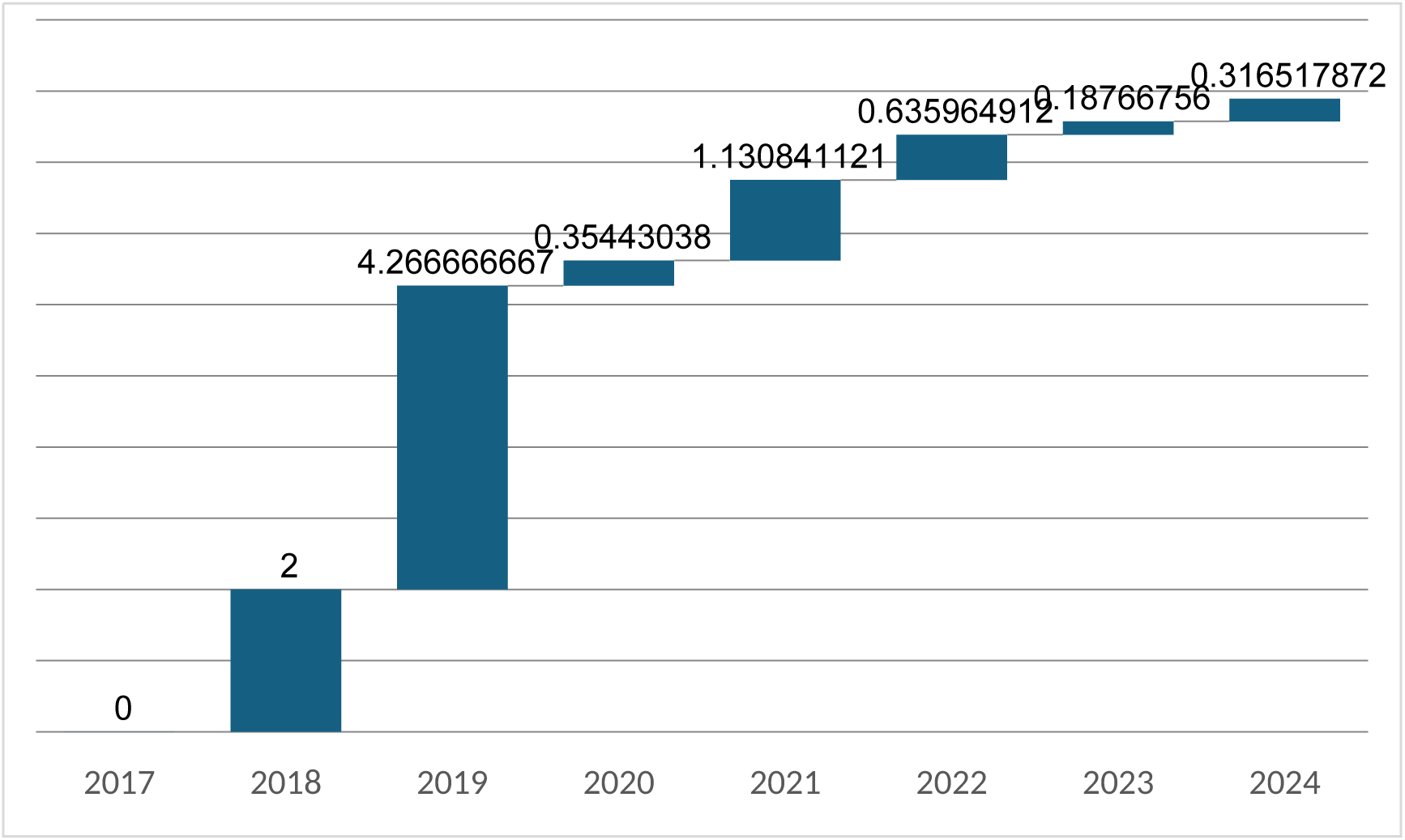
Year-on-year growth (%) of Chronic Disease Medicine Program expenditure 2017-2024. Source: https://nha.moh.gov.ge/ge/c/statics

### Persistent Challenges

Despite these achievements, the program faces several unresolved challenges that limit its effectiveness. Early program design prioritized cost containment over patient-centered care, with medicines selected based on price rather than clinical efficacy or patient/doctor preferences, as also seen in other settings (Bigdeli et al., 2015). This cost-driven approach has also impacted the adoption of certain essential treatments— a study by Murphy et al. found that limited adoption of fixed-dose combinations (FDCs) was due to cost concerns, lack of provider awareness, and procurement challenges, despite their inclusion in the WHO Essential Medicines List(Murphy et al., 2025). The exclusion of combination therapies—despite their alignment with clinical guidelines-also contributed to low uptake and patient dissatisfaction during the early days. Political influences have often superseded technical rationale in decision-making, further undermining program effectiveness, which is not only relevant to Georgia (Kieslich et al., 2016; Lin, 2022). Going forward would require addressing these shortcomings and integrating cost-benefit or health technology assessments (HTAs) into medicine selection processes to ensure better alignment with population health needs and more technically sound allocative decisions. However, the lack of expertise in health economics and the absence of HTA structures will continue to impose barriers unless timely investments are made for capacity strengthening and necessary institutional development (Castro et al., 2020).

Governance and capacity constraints remain significant barriers that need attention. Governance challenges related to leadership and role distribution ambiguities among MoH departments and program stakeholders persist. The lack of a clear and truly participatory leadership structure has led to bureaucratic inefficiencies, with political priorities often overriding technical considerations. Without well-defined roles, accountability is weakened, hindering efforts to address challenges and ensure efficient service provision. Next, the challenges of transparency and public accountability are also areas of concern. While the NHA publishes annual reports, they are not sufficiently granular, easily accessible and/or comprehensible to the general public, constraining transparency and public scrutiny. Therefore, improved governance, including a more explicit definition of decision-making roles and processes with due attention to technical soundness and enhanced transparency, is crucial to ensure equitable access and resource optimization (Haffner et al., 2018).

The absence of a robust digital monitoring system, which currently demands significant analytical expertise and time investments, limits the MoH’s ability to further enhance program management, including identified fiscal risks, particularly following the removal of reimbursement limits. Additionally, insufficient technical expertise in budget forecasting and procurement planning, also noted by other authors (Murphy et al., 2025) has contributed to inefficiencies in resource allocation and is expected to continue challenging the CDMP implementation in the future, unless capacity constraints are timely addressed.

Inadequate public awareness also continues to hinder program uptake. Limited public awareness campaigns might have left many potential beneficiaries unaware of their entitlements, particularly in rural and non-Georgian-speaking regions, leading to regional disparities in coverage (Figure 4). Therefore, targeted, multilingual campaigns leveraging diverse communication platforms, and effective engagement/enhancement of the role of PHC in patient education, are essential to improving awareness and participation, particularly in underserved regions (Pinto et al., 2022; Ranjbar et al., 2017).

**Figure 4.**
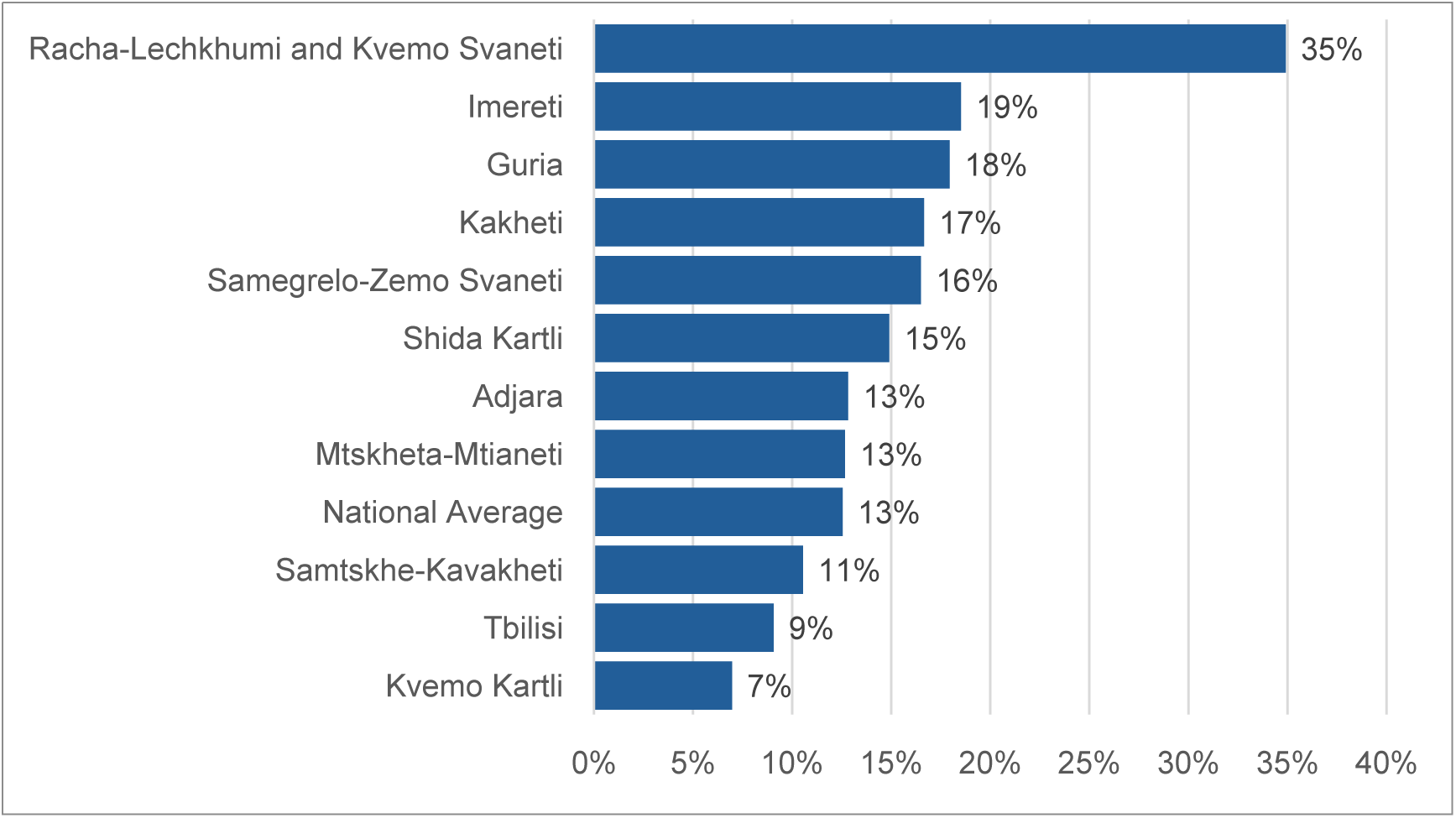
Chronic Disease Medicine Program regional coverage as a percentage of adult population. Source: Reprinted from “Financial burden of healthcare expenditures in Georgia: Challenges and perspectives for the population” by G. Gotsadze, T. Gorgodze, A. Tsuladze, & N. Kotrikadze, 2025, ResearchGate (https://shorturl.at/DKcQd). Copyright 2025 by the authors. DOI: 10.13140/RG.2.2.24611.77602.

PHC providers still play a rather limited role in program implementation. Sometimes, provider behavior highlights the absence of ethical safeguards; they “deny” patients the opportunity to benefit from the program. Observations suggest that unethical pharmaceutical industry practices prevent patients from accessing affordable medications and push them towards more expensive options even when state-funded alternatives are available (Batool et al., 2024; Sanwal et al., 2025; Siddiqui & Siddiqui, 2024). Inadequate PHC engagement and, at times, adverse prescription behavior further constrain CDMP effectiveness. This may stem from inadequate awareness, adverse (unethical) financial incentives and/or a lack of trust in government-funded initiatives. Therefore, further research focused on understanding the incentives influencing PHC provider and pharmacy behaviors and addressing uncovered shortcomings would be crucial for improving program uptake and ensuring long-term sustainability. Meanwhile, it is necessary to strengthen the role of PHC providers through training, incentives, and clear communication to optimize program promotion and patient education and enhance prescription practices (Ez et al., 2010; WHO Regional Office for Europe, 2023). This could also help increase patient awareness about the benefits of the program.

## 6. Study Limitations

This study has certain limitations that should be considered when interpreting the findings. Firstly, the number of respondents interviewed was limited, potentially affecting the generalizability of the findings. Next, the exclusion of provider and patient perspectives and input from pharmaceutical companies or their associations could impose limitations. However, the research team could not include data from these individuals or organizations due to resource and time constraints. Finally, the qualitative nature of the study and the reliance on interview data may introduce some degree of subjectivity and bias in data interpretation, but this weakness was partially addressed with the help of triangulation across primary and secondary (qualitative and quantitative) sources.

## 7. Conclusions

The lessons from Georgia’s CDMP broadly apply to other LMICs striving to design and implement pharmaceutical benefit programs necessary to meet UHC goals of enhancing service use and financial protection. Building on the critical insight that a well-functioning health system is essential for delivering NCD medicines under a UHC scheme, this study emphasizes that addressing inequities in access to medicines requires more than just policy ambition—it demands a comprehensive and integrated approach. Achieving equitable access hinges on leveraging existing capacity for patient-centric program design, effective implementation, strong management, and sustainable financial planning. Without robust governance, continuous capacity-building, and well-functioning monitoring systems and public engagement, even the most well-intentioned policies risk falling short. Strengthening these pillars could ensure that health system components are not only aligned but also resilient enough to deliver sustainable and equitable access to essential medicines.

The CDMP represents a significant step towards reducing the financial burden of chronic diseases in Georgia. However, the program’s sustainability and impact largely depend on addressing persistent design, governance, and public engagement challenges. By prioritizing evidence-based decision-making, strengthening capacity, and fostering equitable access, Georgia can further enhance the effectiveness of the CDMP and contribute to global efforts to achieve UHC.

## Data Availability

All data produced in the present work are contained in the manuscript

## List of Abbreviations

ATM: Access to Medicines
CDMP: Chronic Disease Medication Program
ERP: External Reference Pricing
GDP: Gross Domestic Product
GDP Standards: Good Distribution Practice Standards
HIS: Health Information System
LMICs: Low- and Middle-Income Countries
MoF: Ministry of Finance
MoH: Ministry of Health
NCDs: Non-Communicable Diseases
NCDC: National Center for Disease Control and Public Health
NHA: National Health Agency
OECD: Organization for Economic Co-operation and Development
OOP: Out-of-Pocket (Payments)
PHC: Primary Health Care
SDG: Sustainable Development Goals
STEPs: STEP-wise Approach to NCD Surveillance
UHC: Universal Health Coverage
UHCP: Universal Health Care Program
USAID: United States Agency for International Development

## Authors’ contributions

A.T., A.Z., G.G. and N.K. conceptualized the paper and selected the conceptual framework for the study. A.T. collaborated with N.K. to develop the methodological approach to address the research objectives, conduct the interviews, and analyze the data. J.S. contributed to the literature search and helped structure the introduction section. A.T., N.K. and G.G. played a key role in interpreting the findings and drafting the results section. A.T., K.Z. and G.G. Contributed to the discussion section of the paper. A.Z. and G.G. reviewed and verified the conceptual framework analytical methods, providing critical oversight throughout the study. All authors contributed to drafting, revising, and finalizing the manuscript and approved the final version for submission.

## Funding

This study was funded by the Shota Rustaveli National Science Foundation of Georgia under grant number FR-22-7764. The sponsors were not involved in the study’s design, data collection, analysis, interpretation of results, or the decision to publish.

## Ethics approval and consent to participate

Ethical approval was obtained from the Health Research Union’s ethical committee (protocol #2024-01, approved 04/03/2024). Prior to interviews, all participants provided written informed consent, which included assurances of confidentiality, voluntary participation, and the right to withdraw at any time. The consent forms, available in simplified Georgian, were explained to ensure participant comprehension, and any questions or concerns were addressed before signing.

## Declaration of generative AI and AI-assisted technologies in the writing process

During the preparation of this work the authors used ChatGPT (OpenAI) in order to improve the readability and language of the manuscript. After using this tool, the authors reviewed and edited the content as needed and take full responsibility for the content of the published article

## Footnotes

1 The committee, which operates within the NHA and includes members from both the NHA and the MoH’s policy department, is responsible for reviewing medicine list modification requests submitted to the Ministry

2 There are 12 self-governing cities in the country and 59 self-governing communities/municipalities in Georgia

3 Since 2016, an electronic prescription system has been introduced. From 2022 onward, healthcare providers or individuals with the right to practice independent medical activities are required to prescribe pharmaceutical products exclusively in electronic form.

## Appendix A

**Figure 5.**
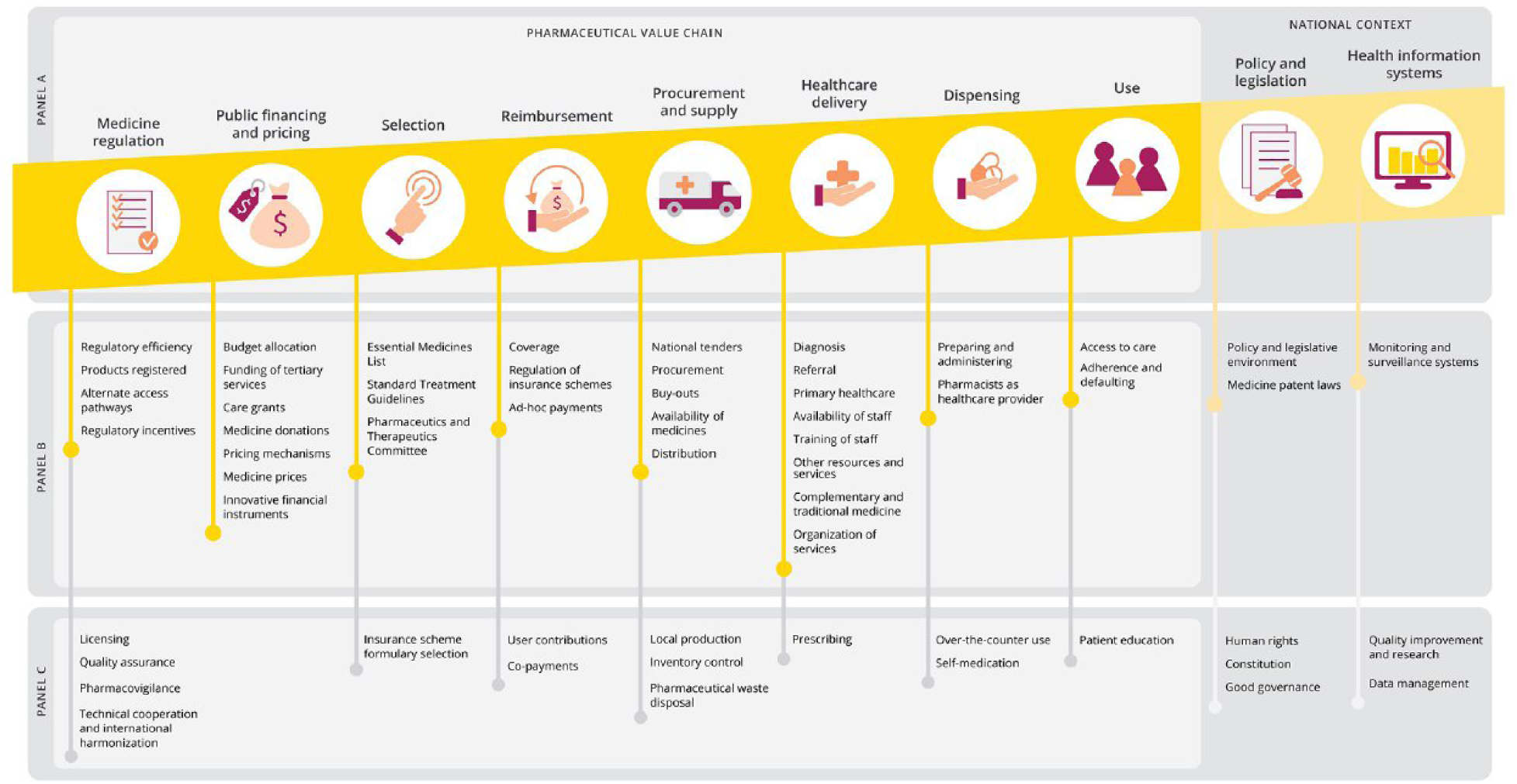
Conceptual framework for qualitative study tool design.

## Appendix B

**Table 2.** Code tree.

| Code Name | Description |
| --- | --- |
| Context | Examines contextual factors that positively or negatively influence the program's performance. |
| Coverage decisions - which population groups to cover | Analysis of the process for selecting beneficiary groups and evaluates respondents' perceptions of the program's coverage scope. |
| Healthcare delivery and dispensing medicines | Explores the challenges encountered by program beneficiaries and administrators in delivering healthcare services and dispensing medicines. |
| Administrative barriers | Identifies bureaucratic and organizational obstacles affecting program implementation. |
| Geographical barriers | Explores access challenges stemming from physical distance or inadequate infrastructure. |
| Financial access barriers | Assesses economic challenges preventing beneficiaries from accessing healthcare services. |
| Dispensing of medicines and its evolution | Reviews provider and pharmacy-level dispensing practices and its progression over time, including medicine stock management. |
| Dispensing practices among providers | Highlights dispensing practices observed among healthcare providers. |
| Dispensing practices among pharmacists | Focuses on dispensing behaviours among pharmacists. |
| PHC system | Evaluates the role and effectiveness of the PHC system in supporting program execution. |
| Inventory control & stock management | Explores challenges in medicine stock management at healthcare facilities and solutions adopted. |
| Monitoring arrangements | Details program monitoring mechanisms, including performance |

| **Code Name** | **Description** |
| --- | --- |
| for program performance | indicators and targets. |
| Outcomes | Covers program outcomes such as coverage and uptake |
| Coverage Gaps | Addresses issues of limited population reach. |
| Low program uptake | Identifies factors contributing to low program utilization among beneficiaries. |
| Procurement and Supply | Analyzes procurement systems, distribution arrangements, and tendering mechanisms over time. |
| Availability of medicines | Discusses challenges in maintaining a consistent supply of medicines. |
| Distribution arrangements for medicines and its evolution | Tracks changes in medicine distribution processes. |
| National procurement arrangements and its evolution | Details the historical development of national procurement and tendering systems. |
| Public Financing and Pricing | Covers budget calculation steps, decision-making entities, data use for budgeting, and price management mechanisms. |
| Budget determination and allocation | Explores steps for budget determination, decision-making entities, and data-driven budgeting processes. |
| Medicine pricing mechanisms | Analyzes strategies for managing medicine prices, including reference pricing. |
| Reimbursement | Covers co-payment history, reimbursement arrangements and their evolution, and reimbursement limits. |
| Co-payment arrangements | Reviews the history of beneficiary cost-sharing mechanisms. |
| Reimbursement | Explores changes in reimbursement arrangements and limits over |
| arrangements | time. |
| Determination of reimbursement limits | Highlights caps set for financial risk protection under UHC policies. |
| Selection-Medicines Selection | Examines the criteria, decision-makers, and processes involved in medicine selection. |
| Selection process | Examines the criteria, decision-makers, and processes involved in medicine selection. |
| External Contributions | Evaluates the role of external stakeholders in the selection process. |
| Committee functioning | Analyzes the effectiveness of selection committees in decision-making. |
| Program Utilization | Covers patients' awareness and preferences regarding the program, medicines formulation demand, and effectiveness of awareness-raising activities |
| Population preferences | Assesses demand for specific medicine formulations/molecules among beneficiaries. |
| Patient awareness | Measures patient awareness of program benefits and the effectiveness of outreach efforts. |
| Recommendations | Summarizes respondents' suggestions for improving program performance and addressing identified challenges. |

## Notes

### Competing Interest Statement

The authors have declared no competing interest.

### Author Declarations

Ethics committee/IRB of Health Research Union gave ethical approval for this work. Protocol #2024-01, approved 04/03/2024

